# Ultrametric model for covid-19 dynamics: an attempt to explain slow approaching herd immunity in Sweden

**DOI:** 10.1101/2020.07.04.20146209

**Authors:** Andrei Khrennikov

## Abstract

We present a mathematical model of infection dynamics that might explain slower approaching the herd immunity during the covid-19 epidemy in Sweden than it was predicted by a variety of other models; see graphs Fig. 2. The new model takes into account the hierarchic structure of social clusters in the human society. We apply the well developed theory of random walk on the energy landscapes represented mathematically with ultrametric spaces. This theory was created for applications to spin glasses and protein dynamics. To move from one social cluster (valley) to another, the virus (its carrier) should cross a social barrier between them. The magnitude of a barrier depends on the number of social hierarchy’s levels composing this barrier. As the most appropriate for the recent situation in Sweden, we consider linearly increasing (with respect to hierarchy’s levels) barriers. This structure of barriers matches with a rather soft regulations imposed in Sweden in March 2020. In this model, the infection spreads rather easily inside a social cluster (say working collective), but jumps to other clusters are constrained by social barriers. This model’s feature matches with the real situation during the covid-19 epidemy, with its cluster spreading structure. Clusters need not be determined solely geographically, they are based on a number of hierarchically ordered social coordinates. The model differs crucially from the standard mathematical models of spread of disease, such as the SIR-model. In particular, our model describes such a specialty of spread of covid-19 virus as the presence of “super-spreaders” who by performing a kind of random walk on a hierarchic landscape of social clusters spreads infection. In future, this model will be completed by adding the SIR-type counterpart. But, the latter is not a specialty of covid-19 spreading.

## 1 Introduction

The dynamics of propagation of the covid-19 pandemy has many unusual features that were not reflected by classical matematical modeling of epidemic dynamics (see, e.g., [1]–[4]). In spite the tremendous efforts [5]–[11], we still do not have a mathematical model describing adequately this dynamics. In this complex situation, one can consider models reflecting only some *specialties of propagation of covid-19 infection* throughout the human society. This paper is a step in this direction. We want *to explain essentially slower than expected approaching the herd immunity*^1^ *in Swedish population* (see, e.g., [12]–[14] for reports from Public Health Institute of Sweden, [7]–[9] for attempts of mathematical modeling and [32]–[36] for reports from massmedia).^2^ Our model is is based on the advanced mathematics, theory of random walks on hierarchic structures. We use the results of the well known paper [19] on random walk on *p*-adic trees, the simplest hierarchic structures endowed with ultrametric.

The main output that can be interesting say for epidemiologists are the graphs presented in section 4 showing *t*^−*a*^ dynamics of probability to become infected in clustered society in long run of an epidemy; so to say *at the phase of approaching of some immunity level*. The model differs crucially from the standard models of spread of disease, such as the SIR-model [4]. Besides explaining slow growth of immunity, our model describes such a specialty of spread of covid-19 virus as the presence of “super-spreaders” who by performing a kind of random walk on a hierarchic landscape of social clusters spreads infection (section 5). In future, the model will be completed by adding the SIR-type counterpart. But, the latter is not a specialty of covid-19 spreading. We hope that this paper may attract attention of experts in epidemiology and sociology (the model explores the structure of social connections in the society) who may stimulate its further development.

Our basic assumption (that, in fact, led to consideration of the ultrametric mathematical model) is that infection’s distribution in the society has the structure of disjoint social clusters.^3^ Such a cluster can be a collective of some enterprise or a state department, say clerks of community office of some town or the personal of some hospital. Inside such a cluster people still have relatively high degree of social connections (see appendix 2). However, even the mild isolation policy of Swedish authorities erected sufficiently high *barriers between clusters*, because people terminated many sorts of social contacts. In particular, the government did not recommend travels inside the country or abroad. (They were not strictly forbidden, but the majority of citizens followed these recommendations.) Another characteristic feature of the model is that connections between social clusters that can be used by the virus to spread have *the hierarchic structure*. For example, the community office has higher level in the social hierarchy than the working place. Any worker can have some things to do with community even during the period of epidemy.

Thus the basic assumption of our model is the following one:

**Assumption:** *Infection distribution in population has the hierarchic social cluster structure*.

Starting with this assumption, we design a mathematical model implying

**Consequence 1**. *Slower than predicted approaching the herd immunity*.

**Consequence 2**. *Slow decay of epidemy*.

The problem of approaching the herd immunity is especially important in the light of Swedish experience and its consequences for states’ policies in preparation to the second wave of covid-19 or other similar viruses. The herd immunity is mathematically formalized through the probability for a person to become infected at time *t*, see Fig. **??** (section 4).

To model cluster dynamics, we use random walk on ultrametric spaces [19]. It was widely used in studied in physics and microbiology, see [19], [20]–[25] and references herein. Geometrically ultrametric spaces have the treelike structure. The simplest trees are homogeneous trees with the fixed number of branches *p* leaving each vertex, *p*-adic trees.

Random walks on trees describe dynamics on *energy landscapes*. There are given energy barriers Δ_*m*_ separating valleys, movement from one valley to another valley is constrained by necessity to jump over a barrier between them.

In our model of the covid-19 epidemy, the virus (or its carrier) randomly walks in socially clustered society. In our model, it starts just from a single social cluster (this assumption is used for mathematical simplicity); it spreads relatively easily inside any cluster, but to approach other social clusters it should “jump over social barriers”. Dynamics depends heavily on two parameters: on the magnitudes of the social barriers (the type of their growth between different levels of hierarchy) and on the social analog of temperature. The magnitude of a social barrier depends only on the number of hierarchy levels composing it, but not on a social cluster. This is the very important constraint on the population, it should be sufficiently homogeneous with respect to the magnitudes of social barriers.

The configuration space of dynamics is the tree of social connections between people. In epidemy modeling, it is natural to assume the presence of the hierarchic structure in social clustering of people, by ranging basic social parameters coupled to infection. This representation of individuals, as *vectors of hierarchically ordered social coordinates* have been already used by the author and his collaborators in a series of studies in cognition, psychology, and sociology [26]–[31].

Just before submission of this preprint, I discovered the recent paper of Britton et al. [15]. One of its authors, Britton, initiated intensive mathematical modeling of spreading covid-19 in Sweden and, in particular, he is famous in Sweden for his prognoses of approaching the herd immunity in Swedish population [7]–[9]. However, his prognoses did not match with the real dynamics of epidemy in Sweden. In the new modeling [15], Britton et al. started to take into account heterogeneity of population (cf. [16]–[18]). This is a step towards coupling with our model. Before to discuss similarities and differences of two models in more detail, I should study article [15] more carefully.

## 2 Social trees

We represent the human society as a system of hierarchically coupled disjoint clusters. There are many ways of creation of such representa-tions. We present one of these possibilities that was used in my works of ultrametric modeling in cognition and sociology (see, e.g., [26]–[30]). The treelike representation of *social types* is based on selection of hierarchically ordered social factors enumerated as *m* = 0, 1, 2, 3, …; factor *m* = 0 is the most important, *m* = 1 is less important and so on. A social type is represented by a vector

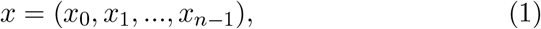

where its coordinates *x*_*m*_ take (typically) discrete values quantifying the *m*-th factor. In the simplest case, *x*_*m*_ takes two values, “yes”/”no”, or 1/0. We call numbers (*x*_*m*_) *social coordinates*. The vector representation of social types and individuals is widely used in sociology and psychology. The main distinguishing feature of our model is endowing the space of vectors with the special metric reflecting the hierarchic structure corresponding the order of social factors (see, e.g., [26]–[30] on application to cognition and psychology). The space of all vectors of the form (4) is called the hierarchic social space.

Since in the covid-19 situation the state plays the crucial role as the policy determining organ, it is natural to select *m* = 0 as the state that population is under consideration. However, since the majority of states selected the lock down policy that was not oriented towards approaching the herd immunity, we restrict consideration to the Swedish population and use *m* = 0 for the next basic social factor, namely, for individual’s age, then say *m* = 1 for the presence of one chronic diseases, *m* = 2 for gender, *m* = 3 for race, *m* = 4 for the town of location, *m* = 6 for profession, *m* = 7 for the level of social activity, *m* = 5 the district in the town, *m* = 8 for the number of children, and so on. We understand that such ranking of the basic social factors related to the covid-19 epidemy may be naive and incomplete. The contribution of sociologists, psychologists, and epidemiologists can improve the present model essentially.

For mathematical simplicity, we consider *p*-adic coordinates, *x*_*m*_ = 0, 1, …, *p* − 1, where *p >* 1 is a prime number. The space of all such vectors denote by the symbol *Z*_*p*;*n*_ (*p* is fixed). For analytical computations, it is convenient to represent elements of this space by natural numbers

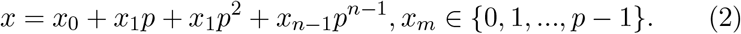

Thus the points of the hierarchic social space can be represented by natural numbers, *Z*_*p*;*n*_ = {0, …, *p*^*n*^ −1}. The number of its points grows exponentially with the number of social coordinates *n*.

We now turn to the definition of a metric on *Z*_*p*;*n*_. Consider two social vectors *x* = (*x*_1_, …, *x*_*n*−1_) and *y* = (*y*_1_, …, *y*_*n*−1_). Let their first *k* coordinates are equal, *x*_0_ = *y*_0_, …, *x*_*k*−1_ = *y*_*k*−1_, but the *k*th coordinates are different, *x*_*k*_ *≠ y*_*k*_. Then the hierarchic social distance between these social types, *d*(*x, y*) = *n*−*k*. The first social coordinates are the most important: longer the common initial segment of vectors corresponds to closer social type, increase of *k* implies decrease of distance between two social types. For example, let *k* = *n* − 1, i.e., two points differ only in the last coordinate, then *d*(*x, y*) = 1. This is the minimal possible distance in *Z*_*p*;*n*_. (The coordinate *x*_*n*_ has the minimal degree of importance.) If the vectors differ already by the first coordinate, i.e., *x*_0_ *≠ y*_0_, then *d*(*x, y*) = *n*. This is the maximal possible distance between points in space *Z*_*p*;*n*_. Distance *d* is ultrametric, it satisfies *the strong triangle inequality:*

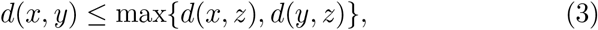

for any triple of points *x, y, z* ∈ *Z*_*p*;*n*_. Here in each triangle, the third side is less or equal not only to the sum of two other sides (as usual), but even to their maximum.

As usual in a metric space we can introduce balls, *B*_*N*_ (*a*) = {*x* ∈ *Z*_*p*;*n*_ : *d*(*a, x*) ≤ *N*}, where *N* = 1, …, *n*, and *a* = (*a*_0_, ..,, *a*_*n*−1_) is some point in *Z*_*p*;*n*_, ball’s center. In an ultrametric space, *any two balls are either disjoint or one is contained in another and any point of a ball can be selected as its center*.

For our modeling, it is important that the space *Z*_*p*;*n*_ can be split into disjoint social clusters. (As we shall see soon, these clusters are, in fact, balls.) Each cluster is determined by fixing a few first (the most important) social coordinates,

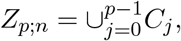

where *C*_*j*_ = {*x* : *x*_0_ = *j*}. This cluster representation corresponds to the first level of social hierarchy, we distinguish points by their most important coordinate. Each of clusters *C*_*j*_ can be represented similarly as

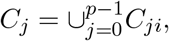

where *C*_*ji*_ = {*x* : *x*_0_ = *j, x*_1_ = *i*} are clusters of the deeper hierarchic level and so on, up to the single-point clusters corresponding to fixing all social coordinates.

Clusters are, in fact, ultrametric balls:

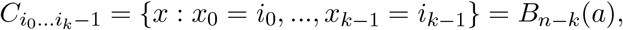

where *a* is any point of the form *a*_0_ = *i*_0_, …, *a*_*k*−1_ = *i*_*k*−1_ and arbitrary coordinates *a*_*j*_, *j* = *k*, …, *n* − 1.

Geometrically space *Z*_*p*;*n*_ is represented as a homogeneous tree with *p* branches leaving each vertex. Cluster is a bunch of branches having the common root. By extending this root we split the cluster into subclusters.

Now we consider the procedure of extension of a social tree by adding new social coordinates, so from tree *Z*_*p*;*n*_ to tree *Z*_*p*;*N*_, where *N > n*. As the result of such an extension, each point of social space *Z*_*p*;*n*_ becomes a social cluster in social space *Z*_*p*;*N*_. In principle, it is impossible to determine a social type by fixing any finite number of social coordinates. Hence, We have to consider infinite sequences of coordinates:

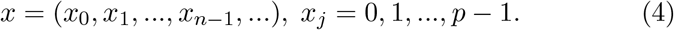

Denote the space of such sequences by the symbol *Z*_*p*_. This is the complete hierarchic social space. Points of finite trees represent social clusters.

## 3 Dynamics

Consider a tree with *n* levels of social hierarchy, *Z*_*p*;*n*_. Suppose that initially infected people are concentrated just in one social cluster represented by a single point *x* ∈ *Z*_*p*;*n*_. Consider the initial probability for a person in this cluster to become infected, *P* (*y*, 0) = *δ*(*y* − *x*).

The virus plays the role of a system moving through barriers in models of dynamics on energy landscapes (see [19], [20]–[25] and references herein). In our case, these are social barriers between social clusters of population. The virus performs a complex random walk motion inside each social cluster moving in its sub-clusters, goes out of it and spreads through the whole population, sometimes the virus comes back to the original cluster from other social clusters that have been infected from this initial source of infection, and so on. During this motion the virus should cross numerous social barriers.

Instead of virus walking through the social tree, we can consider a person. A person of the social type *x* = (*x*_1_, …, *x*_*n*_) can interact with persons of the social type, *y* = (*x*_1_, …, *x*_*k*_ − 1, *y*_*k*_, …, *y*_*n*_), where *x*_*k*_ *≠ y*_*k*_ by crossing the social barrier for *n* − *k* levels of hierarchy. The mtemporal sequence of social contacts of some persons can have a very complicated trajectory, visiting numerous clusters (but the probability of approaching a cluster depends crucially on social barriers).

We are interested in *the relaxation process* that is understood as the evolution of the probability to become infected for a person from the social cluster *x*, i.e., in the time dependence of the quantity *P* (*x, t*).

We do not want to go into details, since the problem of relaxation was well studied in physics and microbiology, see [19], [20]–[25] and references herein. The relaxation regime depends crucially on the barriers’ magnitude, how rapidly they grow up on the way from one cluster to another.

## 4 Random walk on on infinite trees: temporal behavior of probability

As in the previous section, we a random walk on a finite tree. Here we follow the paper of Ogielski [19]. Let us consider a finite tree with *n* levels. Thus there are 2^*n*^ points at the last level. They enumerate the total population: *i* = 0, …, 2^*n*^ − 1.

Let virus encounters a barrier of size Δ_*m*_, in hopping a distance *m* (crossing *m* levels of hierarchy), where Δ_1_ *<* Δ_2_ *<* … *<* Δ_*m*_ *<* It is supposed that barriers Δ_*m*_ are the same for all social clusters, i.e., they depend only on distance, but not on clusters.

The probability to jump over the barrier Δ_*m*_ has the form (up to the normalization constant):

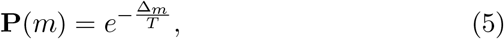

where *T* is model’s parameter; in physics it has the meaning of *temperature* of the environment; we also interpret it similarly, as a kind of *social temperature*, cf. [29]. Probability **P**(*m*) grows with growth of social temperature *T*. For high social temperatures, jumps are easier.

Consider the energy landscape with a uniform barrier Δ, at every branch point; that is, a jump of distance 1 involves surmounting a barrier Δ, of distance 2, a barrier 2Δ, and so on. Hence, barriers linearly grow with distance *m*,

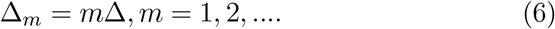

It seems that this type of behavior is the most natural from the viewpoint of social connections during the covid-19 epidemy in Sweden. Barriers are sufficiently high, but they still are not walls as during the rigid quarantine (as say in Italy, France, or Russia). The probability to jump over the barrier Δ_*m*_ has the form (up to the normalization constant):

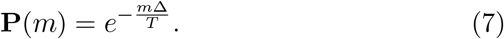

The probability depends on the height Δ of the minimal social barrier, but, for large *m*, its contribution to is not so important.

By using random walk on the tree with *n* levels of hierarchy and approaching *n* → ∞ one can derive the following asymptotic behavior of the relaxation probability:

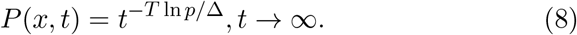

Set *κ* = *T* ln *p/*Δ. If *κ <≪* 1, e.g., the social temperature is low and the primary social barrier Δ is relatively large, then the probability for a person in the original social cluster *x* to become infected decreases rather slowly, see Fig. 1.

**Figure 1:**
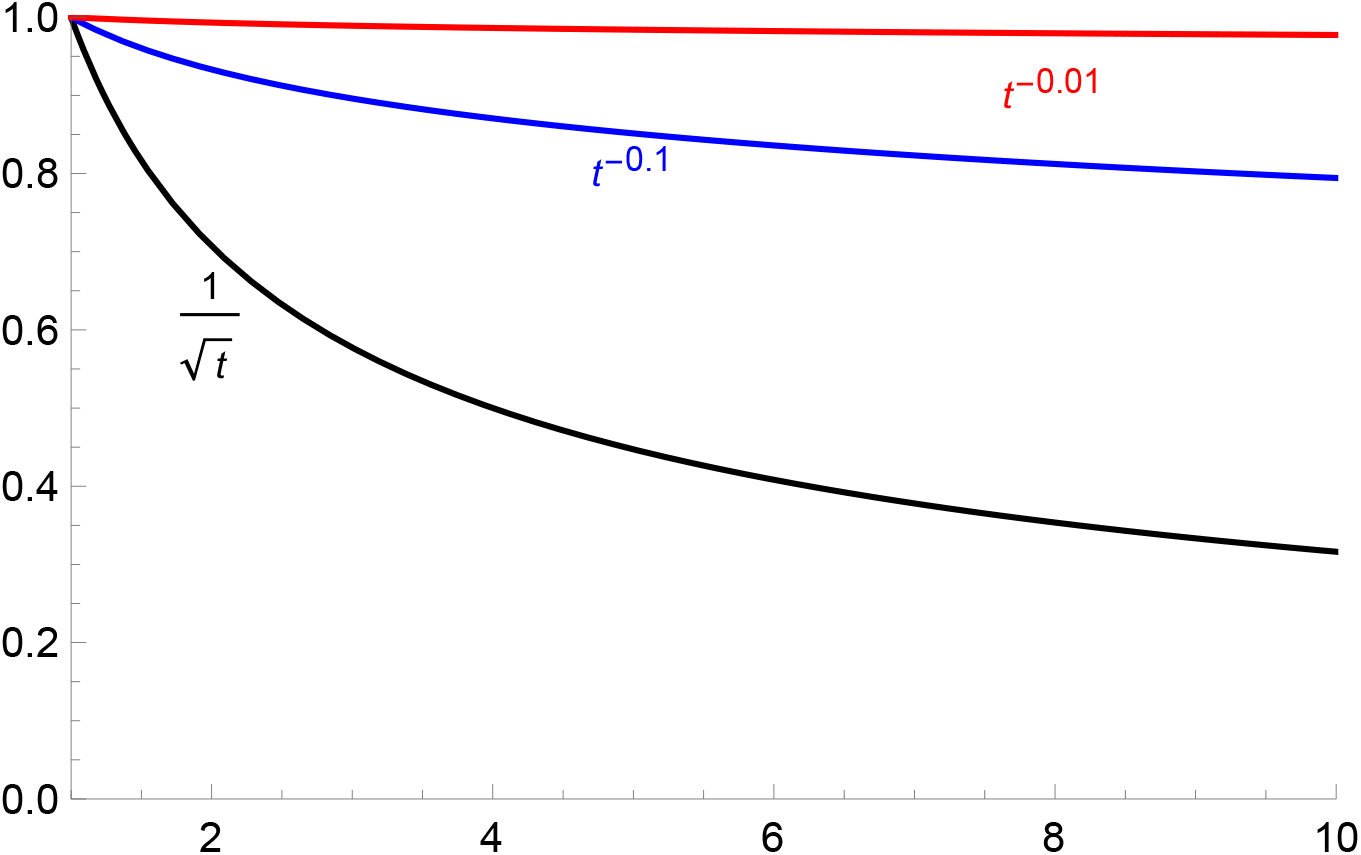
Asymptotic behavior of probability to become infected. (For fixed social temperature *T*, the upper graphs correspond to one-step barrier growth 10 and 100 times, respectively.

Hence immunity against covid-19 increases also slowly, see Fig. 2, as function

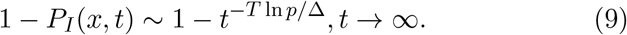

**Figure 2:**
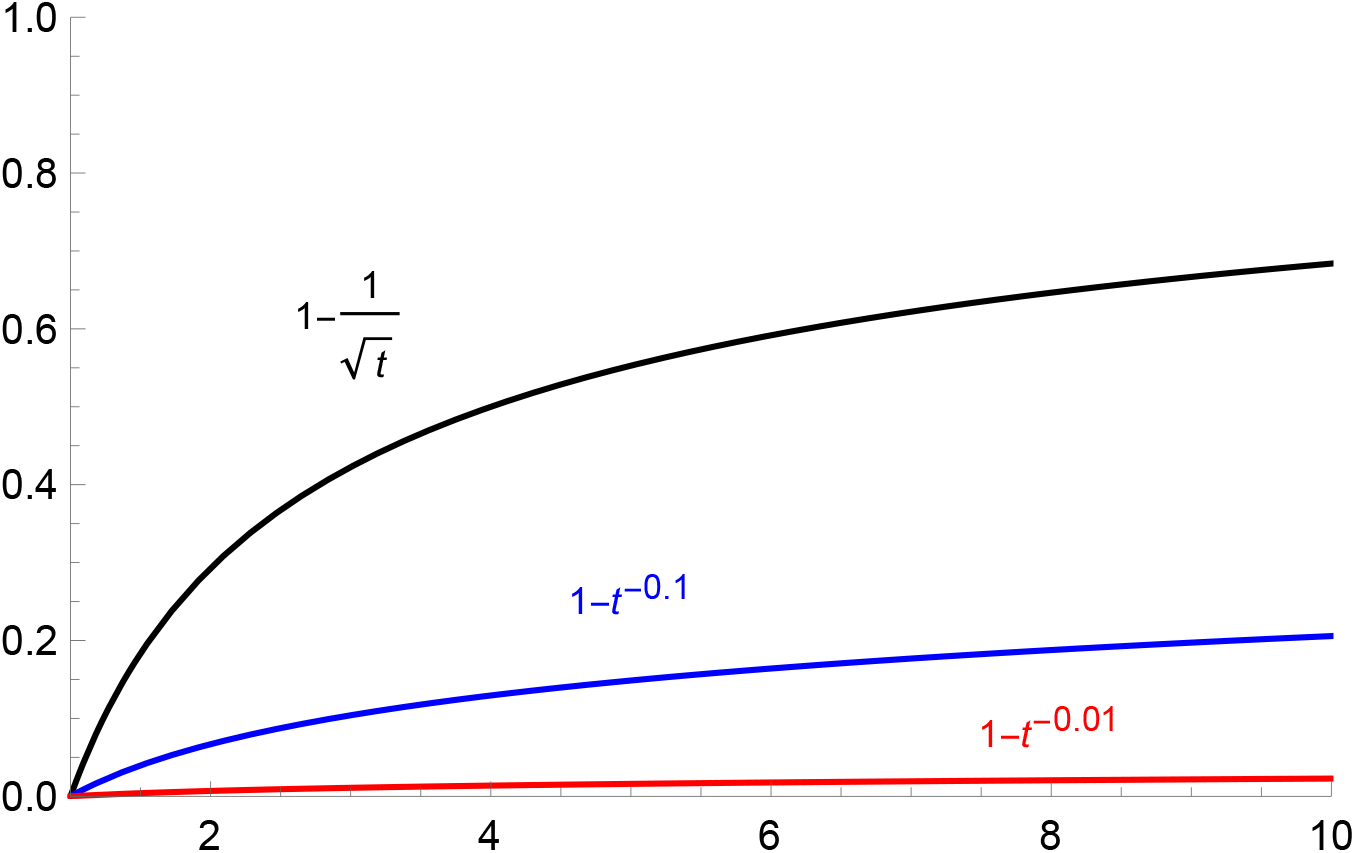
Asymptotic behavior of probability to become immune; increasing of the herd immunity (for fixed social temperature *T*, the upper graphs correspond to one-step barrier growth 10 and 100 times, respectively.

## 5 Average social distance traveled by infection spreader

In our mathematical model, any infection spreader travels through the social tree, he-she visits a few social clusters and infects people in these clusters. Theory of random walks in ultrametric spaces predicts the average social distance for spreader’s travel through clusters, starting at some fixed cluster *x* and jumping to other cluster *y*,

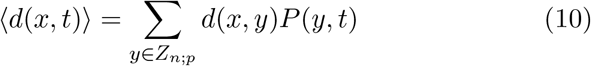

For linearly growing social barriers, and *n* → ∞, the asymptotic behavior has the following form:

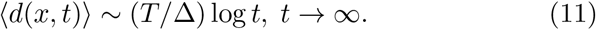

This average distance goes to infinity. (As can be expected, higher social temperature *T* and lower one-step social barrier Δ induce more rapid growth.) Although the log-growth is relatively slow, it, nevertheless, implies very extended spread of the infection. Unsoundness of ⟨ *d*(*x, t*) ⟩ can be associated with the presence of super-spreaders who jump even over high social barriers and spread the virus to social clusters that are far from the original source of infection. We repeat once again that the distance under consideration is in social and not in physical space.

## 6 Concluding remarks

The presented ultarmetric model with random walk dynamics on energy landscapes describes well the social cluster structure of spreading of covid-19. The model was designed to explain mathematically the slow approaching the herd immunity in Swedish population during March-June 2020, essentially slower than it was predicted by all mathematical models.^4^ The model elevates the role of the social dimension of infection spreading comparing with its purely bio-medical dimension. The bio-medical restrictions in Sweden were very soft; in particular, no face-masks, 1,5 meter social distance, public transportation worked more or less as usual (see appendix 2 for details). The most typical infecting took place in working collectives in that social contacts were not occasional, as say in metro, or in a bus, or even in a restaurant during lunch, but close and permanent, on the everyday basis.

We applied to the new area of research, to epidemiology, mathematical theory developed for applications in statistical physics (spin glasses) and microbiology (protein folding): ultrametric random walk describing dynamics on complex energy landscapes with the hierarchic structure of barriers between valleys. The presence of social barriers growing with hierarchy’s levels makes the evolution of epidemy essentially slower than in models which do not take into account the cluster-character of infection spreading. In particular, our model is purely diffusional, cf. with standard SIR-model [4]. We motivate consideration of linearly growing social barriers in Sweden with its soft epidemiological restrictions.

Although the model is very simplified it reflects some features of the covid-19 epidemy, especially its the social cluster character. We hope that our model will stimulate further development of ultrametric epidemiological models that take into account the hierarchic social clustering of population.

## Data Availability

theoretical paper

## Appendix 1: Growth of the herd immunity in Sweden

Our model can be interesting only for Sweden, where the “experiment” for approaching the herd immunity was performed in its clean form, without overshadowing by quarantine. The main concern of Swedish authorities, including Anders Tegnell, is that the dynamics of getting antibodies for covid-19 in Swedish population is essentially slower than it was expected. Since the end of April 2020, Anders Tegnell, the chief state epidemiologist of Sweden, started to announce (see, e.g., [32]) that very soon we, Swedish citizens, will become immune against the virus. This announcement was repeated a few times during May-June 2020, but it still not clear whether it happened or not (see, e.g., [34]–[36]). Moreover, Stockholm university made recently (June 24 []) the update that, in fact, already the level 60% can be considered as the herd immunity state. So, it seems that there is no hope to approach 70% level of immunity.

On June 24, 2020, it was announced [35] that: “A total of 7.3 percent of the blood samples collected from people in Stockholm were positive, 4.2 percent in Skane and 3.7 percent in Västra Götaland. The numbers reflect the situation in early April, as it takes time for the antibodies to develop. The authority’s modeling has previously shown that 26 percent of the population in Stockholm would have been infected on May 1. Tom Britton, professor of mathematical statistics at Stockholm University, comments on these and says that the figures are “clearly lower”, than he thought. “Firstly, I am thinking about the modeling, if there is any mistake that I and the Public Health Authority have made. Then you also wonder if some have been infected but do not have detectable antibodies, for example people who are mildly infected.”

The level of the herd immunity depends on the age group. On June 2, 2020, it was pointed out that for the youngsters up to 19 years old around 8% Andelen med antikroppar mot covid-19 bland yngre upp till 19 år var närmare åtta procent, för åldersgruppen 20-64 låg motsvarande siffra på 6-7 procent. För personer över 65 år låg siffran på cirka tre procent. Det kan finnas flera skäl till att det är lägre nivåer för vuxna och äldre, enligt Karin Tegmark Wisell.

## Footnotes

1 In particular, the mathematical model of covid-19 epidemy dynamics of Tom Britton [7, 8] that was used by Swedish State Health Authority predicted that the herd immunity will be approached already in May.

2 As is well known and widely debated in mass-media, the Swedish government chosen its own way to handle covid-19 epidemy, namely, without any kind of lock-down. The main aim of such a policy is approaching the herd immunity.

3 ”Disjoint” has the meaning of separation in social space. In physical space, clusters can essentially overlap. Social separation does not complete, say of the form of walls. There are social barriers, but a person (virus-carrier) can hope over barriers, with some probability.

4 See appendix 1; finally, the state epidemiologist Anders Tegnell pointed that it seems meaningless to appeal to mathematical models at all, since they do not reflect even approximately the real situation.

## Notes

### Competing Interest Statement

The authors have declared no competing interest.

### Clinical Trial

this is a tehoretical model

### Funding Statement

no funding

